# Comparing survival in urinary tract cancer patients with various second primary malignancies: A population-based study

**DOI:** 10.1101/2022.03.10.22272195

**Authors:** Xiang Gao, Hui Cao, Liang Zhou, Xiaopan Li, Yongbin Zou, Dehua Yu, Jinsong Geng, Haiyi Jia, Yipeng Lv, Wenya Yu, Yichen Chen, Zhaoxin Wang, Jianwei Shi, Hua Jin

**Author notes:** These authors are co-corresponding authors on this work: Zhaoxin Wang; Jianwei Shi; Hua Jin. These authors share first authorship (Xiang Gao, Hui Cao, Liang Zhou).

## Abstract

**Purpose:** We aimed to identify the patterns and combinations of second primary malignancies (SPMs) observed in patients with malignant neoplasms of the urinary tract (MNUT) and to explore the independent risk factors for survival outcomes in these patients.

**Materials and Methods:** We analysed the data of MNUT patients with SPM in 25 hospitals in Shanghai between 2002 and 2015. A life table was used to calculate the survival rates, Kaplan–Meier analysis was used to determine the survival status of MNUT patients, and Cox regression analysis was used to perform multivariate analysis of survival risk factors in MNUT patients with SPM.

**Results:** Among the 154 patients included, the first primary malignancy (PM) most commonly occurred in the bladder (50.65%) and kidney (41.56%), and the SPM most commonly occurred in the lung (22.73%) and stomach (13.64%). The most common combinations included the bladder + lung and bladder + stomach. The Cox regression results showed that age older than 60 years (HR = 2.36 [95% CI 1.30–4.28] vs. age ≤60 years, p = 0.005), TNM 1 stage III+IV disease (HR = 2.19 [95% CI 1.37-4.57] vs. I+ II), p = 0.037), TNM 2 stage III+IV disease (HR = 7.43 [95% CI 1.49-19.68] vs. I + II), p <0.001), and SPM in the lung (HR = 4.36 [95% CI 1.74-18.69], p = 0.047) were associated with a significantly worse cancer-specific survival.

**Conclusion:** The survival of MNUT patients with SPM may be related to the SPM site, first and second PM staging and latency time.

## Introduction

Cancer is the leading cause of death in the world. In 2020, cancer caused nearly 10 million deaths worldwide. The number of new occurrences and deaths of malignant neoplasms of the urinary tract (MNUT) has increased year by year, with figures for bladder and kidney neoplasms reaching 1.005 million and 392,000, respectively(1). Benefiting from the great advances in diagnostic techniques and treatment methods for MNUT, the survival rate of patients is gradually improving(2, 3). Studies have shown that second primary malignancy (SPM) is a serious and potentially fatal long-term complication of cancer patients, which places a heavier burden on the survival of patients with MNUT(4). Therefore, it is important to track and manage the survivors of MNUT and research SPM.

Previous studies have analysed the risk and influencing factors of SPM for specific MNUTs. For example, Beisland, C et al. analysed the multiple primary malignant tumours of 1,425 patients with renal cell carcinoma in Norway and explored the incidence of multiple malignant tumours of renal cell carcinoma and their combined pattern(5). Liu, Y et al. downloaded SPM sample data from the Surveillance, Epidemiology, and End Results (SEER) database, analysed the incidence of SPM in patients with prostate cancer, and further clarified the common sites of SPM and the influencing factors of survival(6). Some scholars have also analysed the survival risk of specific MNUT patients with SPM. For example, Kyo Chul Koo et al. analysed the incidence of SPM in prostate cancer patients and the influencing factors of survival(7). However, to the best of our knowledge, current research focuses more on a specific MNUT and is limited to fewer data, and the overall pattern of MNUT is not yet clear. Moreover, few studies on survival models consider the impact of SPM on the survival of patients with MNUT, and it has been found that the location of malignant tumours is an important prognostic factor for the survival rate of SPM patients.

Therefore, the purpose of this study was to first establish the SPM patterns of MNUTs overall by using measurement data from 25 hospitals in Shanghai, China, and then analyse the survival of common SPM categories in terms of the first MNUT and its SPM site. Finally, we assessed the factors affecting cancer mortality and all-cause mortality in SPM patients.

## Materials and methods

### Data collection

The diagnosis and survival data of MNUT patients with SPM in this study were collected from 25 hospitals in Shanghai. The assessment and examination of data quality was based on the China Cancer Registration Guidelines and the International Agency for Research on Cancer/International Association for the Registration of Cancer data quality standards. Relevant population data were provided by the Shanghai Bureau of Statistics and the Public Security Bureau.

We analysed the data of MNUT patients with SPM from 25 hospitals in Shanghai, China, between 1 January 2002 and 31 December 2015. In addition, we included patients who were diagnosed with MNUT before 2002 but were subsequently diagnosed with SPM in 2002–2015. After patients provided informed consent, we conducted annual follow-up surveys of patients through home visits or telephone calls according to standard epidemiological procedures to assess the survival rate(8). The follow-up observation continued until the date of death or December 31, 2017, to resolve the 3-year time lag in information collection and data quality control. According to ICD-10, 5 types of urinary tract malignancies (C64-C68) were identified. After screening the first primary malignancy (PM) site of the MNUT and excluding 22 patients with 3 PMs, a total of 154 urinary organ cancer patients with SPM were identified as the subjects of the study (**Figure 1**).

**Figure 1.**
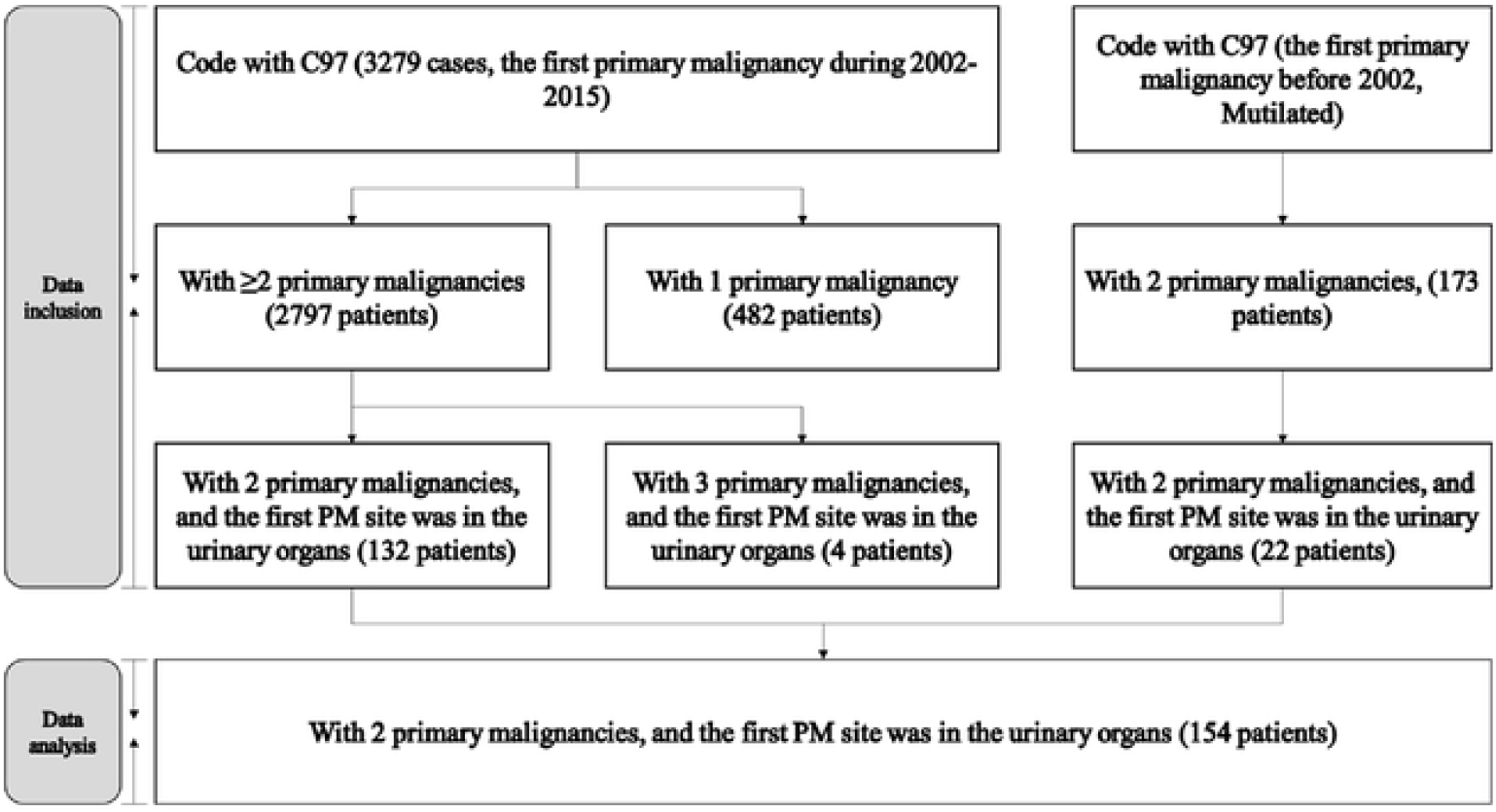
Data source of the included and analysed SPM patients.

### Variables

The following demographical and clinical variables were used in our study: age at diagnosis (patients ≤ 60 years and > 60 years were stratified into the non-older group and older group, respectively(9)), American Joint Committee on Cancer (AJCC) 7th tumour-node-metastasis (TNM) staging classification(10), year of diagnosis, level of hospital, latency (the interval between the first PM diagnosis and the SPM diagnosis(11)) and site of SPM. TNM 1 represents the TNM staging classification of the first PM (MNUT), and TNM 2 represents the TNM staging classification of the SPM.

### Statistical analysis

We used a life table to calculate the 1–5-year survival rate, Kaplan-Meier analysis to determine the survival status of patients with urinary system cancer, and Cox regression analysis for multivariate analysis of survival risk factors for MNUT patients with SPM. A two-sided test was used for analysis, and the statistical software used was SPSS 20.0. P<0.05 was considered to indicate a significant difference.

## Results

### Population and characteristics

Among the 154 MNUT patients with SPM, there were 119 male patients, accounting for 77.27%, and 35 female patients, accounting for 22.73%. From the perspective of age distribution, the patients were mostly over 65 years, accounting for 70.78%. The proportion of patients with TNM 1 and TNM 2 in grade III+IV was higher than the proportion of patients in grade I+II. Most patients were diagnosed with MNUT in tertiary hospitals (76.62%) between 2002 and 2008 (66.88%). In addition, most people were diagnosed with SPM in tertiary hospitals (59.09%) between 2009 and 2015 (81.82%). The latency between the first and second PM diagnoses in most patients was less than 12 months (81.17%). Among the SPM sites of MNUT, lung, stomach and colon were the most common, accounting for 22.73%, 13.64% and 11.69% of the total, respectively (**Table 1**).

**Table 1.** Characteristics of MNUT patients with SPM (n, %)

| Variables | Male | Female | Total |
| --- | --- | --- | --- |
| **Age at SPM diagnosis** |  |  |  |
| ≤60 years | 32(26.89%) | 13(37.14%) | 45(29.22%) |
| >60 years | 87(73.11%) | 22(62.86%) | 109(70.78%) |
| **TNM 1 (MNUT) stage** |  |  |  |
| III+IV | 69(57.98%) | 20(57.14%) | 89(57.79%) |
| I+II | 3(2.52%) | 4(11.43%) | 7(4.55%) |
| Missing | 47(39.50%) | 11(31.43%) | 58(37.66%) |
| **TNM 2 (SPM) stage** |  |  |  |
| III+IV | 32(26.89%) | 10(28.57%) | 42(27.27%) |
| I+II | 17(14.29%) | 12(34.29%) | 29(18.83%) |
| Missing | 70(58.82%) | 13(37.14%) | 83(53.90%) |
| **Time of first PM (MNUT) diagnosis (year)** |  |  |  |
| 2009–2015 | 37(31.09%) | 14(40.00%) | 51(33.12%) |
| 2002–2008 | 82(68.91%) | 21(60.00%) | 103(66.88%) |
| **Time of SPM diagnosis (year)** |  |  |  |
| 2009–2015 | 98(82.35%) | 28(80.00%) | 126(81.82%) |
| 2002–2008 | 21(17.65%) | 7(20.00%) | 28(18.18%) |
| **Hospital level for first PM (MNUT)** |  |  |  |
| Secondary | 28(23.53%) | 8(22.86%) | 36(23.38%) |
| Tertiary | 91(76.47%) | 27(77.14%) | 118(76.62%) |
| **Hospital level for SPM** |  |  |  |
| Secondary | 51(42.86%) | 12(34.29%) | 63(40.91%) |
| Tertiary | 68(57.14%) | 23(65.71%) | 91(59.09%) |
| **Latency between MNUT and SPM** |  |  |  |
| ≤ 12 months | 98(82.35%) | 27(77.14%) | 125(81.17%) |
| > 12 months | 21(17.65%) | 8(22.86%) | 29(18.83%) |
| **SPM site** |  |  |  |
| Lung | 29(24.37%) | 6(17.14%) | 35(22.73%) |
| Stomach | 17(14.29%) | 4(11.43%) | 21(13.64%) |
| Colon | 16(13.45%) | 2(5.71%) | 18(11.69%) |
| Prostate | 11(9.24%) | 0(0.00%) | 11(7.14%) |
| Bladder | 6(5.04%) | 4(11.43%) | 10(6.49%) |
| Others | 40(33.61%) | 19(54.29%) | 59(38.31%) |

Figure 2. shows the distribution of first and second PMs in 154 patients with MNUT. The main sites of MNUT were the bladder (n=78, 50.65%) and kidney (n=64, 41.56%). When the first PM occurred in the bladder, the most common SPM sites were the lungs (n=19, 24.36%), stomach (n=12, 15.38%) and prostate (n=9, 11.54%). When the first PM occurred in the kidney, the most common SPM sites were the lungs (n=12, 18.75%), colon (n=9, 14.06%), bladder (n=8, 12.50%) and stomach (n=8, 12.50%).

The survival rates of first and second PM patients are shown in **Table 2**. Taking the first PM site as the centre, bladder cancer patients had relatively low survival rates, with 1-year, 2-year, 3-year, 4-year and 5-year survival rates of 94.87%, 83.33%, 76.92%, 74.36% and 65.38%, respectively. When the SPM site was the lung, the observed survival rate was the lowest. The 1-year, 2-year, 3-year, 4-year, and 5-year survival rates were 54.29%, 37.14%, 22.86%, 14.29%, and 2.86%, respectively.

**Table 2.** Survival rates of primary malignancy patients based on the MNUT and SPM site during 2002–2015.

| Site | Survival rates (%) |  |  |  |  |  |
| --- | --- | --- | --- | --- | --- | --- |
|  | N | 1-year | 2-year | 3-year | 4-year | 5-year |
| **First primary malignancy** |  |  |  |  |  |  |
| Bladder | 78 | 94.87 | 83.33 | 76.92 | 74.36 | 65.38 |
| Kidney | 64 | 95.31 | 92.19 | 89.06 | 81.25 | 68.75 |
| Others | 12 | 91.67 | 83.33 | 75.00 | 66.67 | 66.67 |
| **Second primary malignancy** |  |  |  |  |  |  |
| Lung | 35 | 54.29 | 37.14 | 22.86 | 14.29 | 2.86 |
| Stomach | 21 | 61.90 | 42.86 | 38.10 | 33.33 | 23.81 |
| Colon | 18 | 83.33 | 72.22 | 55.56 | 16.67 | 5.56 |
| Prostate | 11 | 72.73 | 72.73 | 72.73 | 54.55 | 18.18 |
| Bladder | 10 | 80.00 | 80.00 | 20.00 | 20.00 | 20.00 |
| Others | 59 | 74.58 | 66.10 | 55.93 | 33.90 | 27.12 |

**Figure 2.**
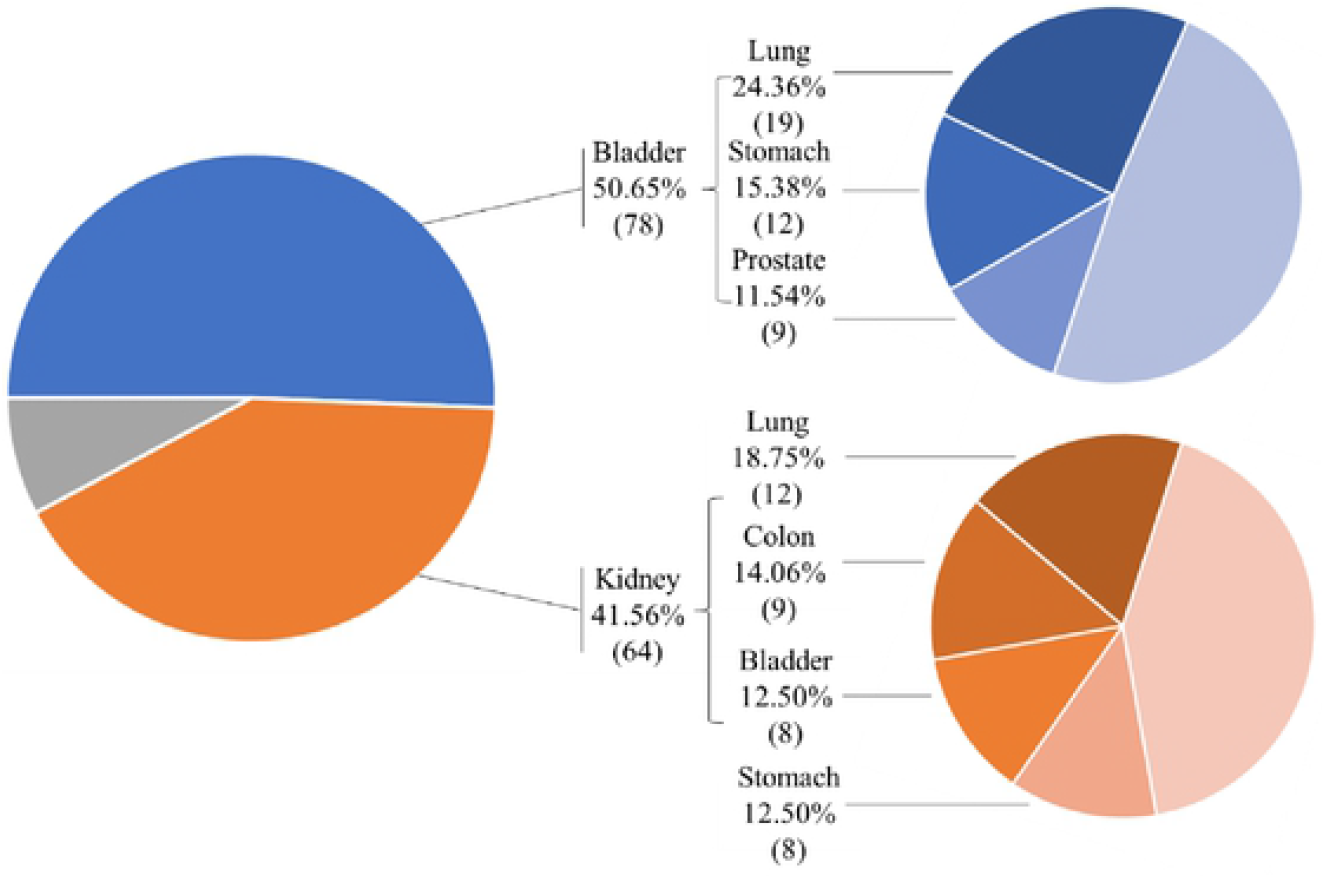
Distribution of MNUT and SPM among all patients.

### Possible risk factors for the survival status of MNUT patients with SPM

Kaplan-Meier analysis showed that there were significant differences in the cancer-specific survival rates of patients with different age groups, TNM levels, and SPM sites (p<0.05) (**Figure 3**). Specifically, patients older than 60 years of age have a higher risk of cancer death than patients younger than 60 years of age. Patients with TNM 2 stage III+IV disease have a higher risk of cancer death than those with TNM 2 stage I+II disease. The risk of cancer death in patients with SPM found in the lung was higher than that in patients with SPM found in the bladder and prostate. Other variables, such as sex, time of diagnosis, hospital level, and time between first and second cancers, had no statistically significant influence on cancer survival. The overall survival rate was the same as the cancer-specific survival rate. Advanced age, TNM stage III+IV disease and lung SPM site were important risk factors for all-cause death.

**Figure 3.**
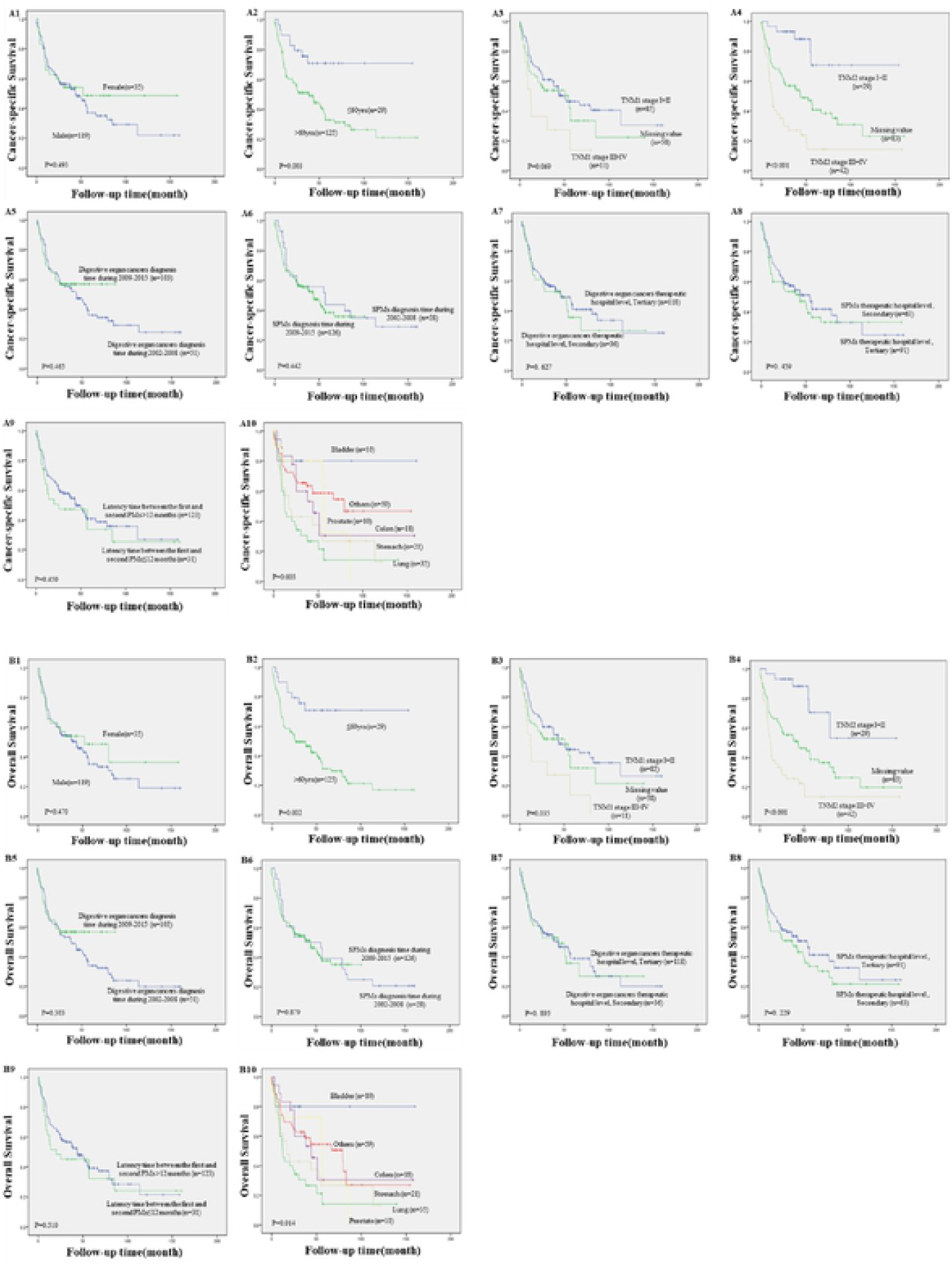
Kaplan-Meier analysis of the survival status and risk factors of the patients. Note: A indicates the cancer-specific survival of the patients, and B indicates the overall survival of the Patients.

After univariate analysis, variables (p<0.3) were selected for Cox regression analysis, and 4 variables were shown to be independent risk factors for cancer survival **(Table 3)**. For cancer-specific survival, patients older than 60 years (compared to patients ≤60 years old) had a 2.36-fold increased risk of death (HR = 2.36 [95% CI 1.30–4.28] vs. age ≤60 years, p = 0.005). TNM 1 III+IV patients had a 2.19-fold increase in the risk of death (HR = 2.19 [95% CI 1.37-4.57] vs. I+ II, p = 0.037). In addition, MNUT patients with TNM 2 stage III+IV disease (compared to patients with TNM 2 stage I+II disease) had a 7.43-fold increase in the risk of death (HR = 7.43 [95% CI 1.49-19.68] vs. I + **II**), p <0.001). It should also be noted that patients with urinary tract malignancies had a significantly higher risk of dying from lung SPM (HR = 4.36 [95% CI 1.74-18.69], p = 0.047). Unlike the cancer-specific survival rate, there was no significant difference in the effect of the site of SPM on the overall survival rate.

**Table 3.**
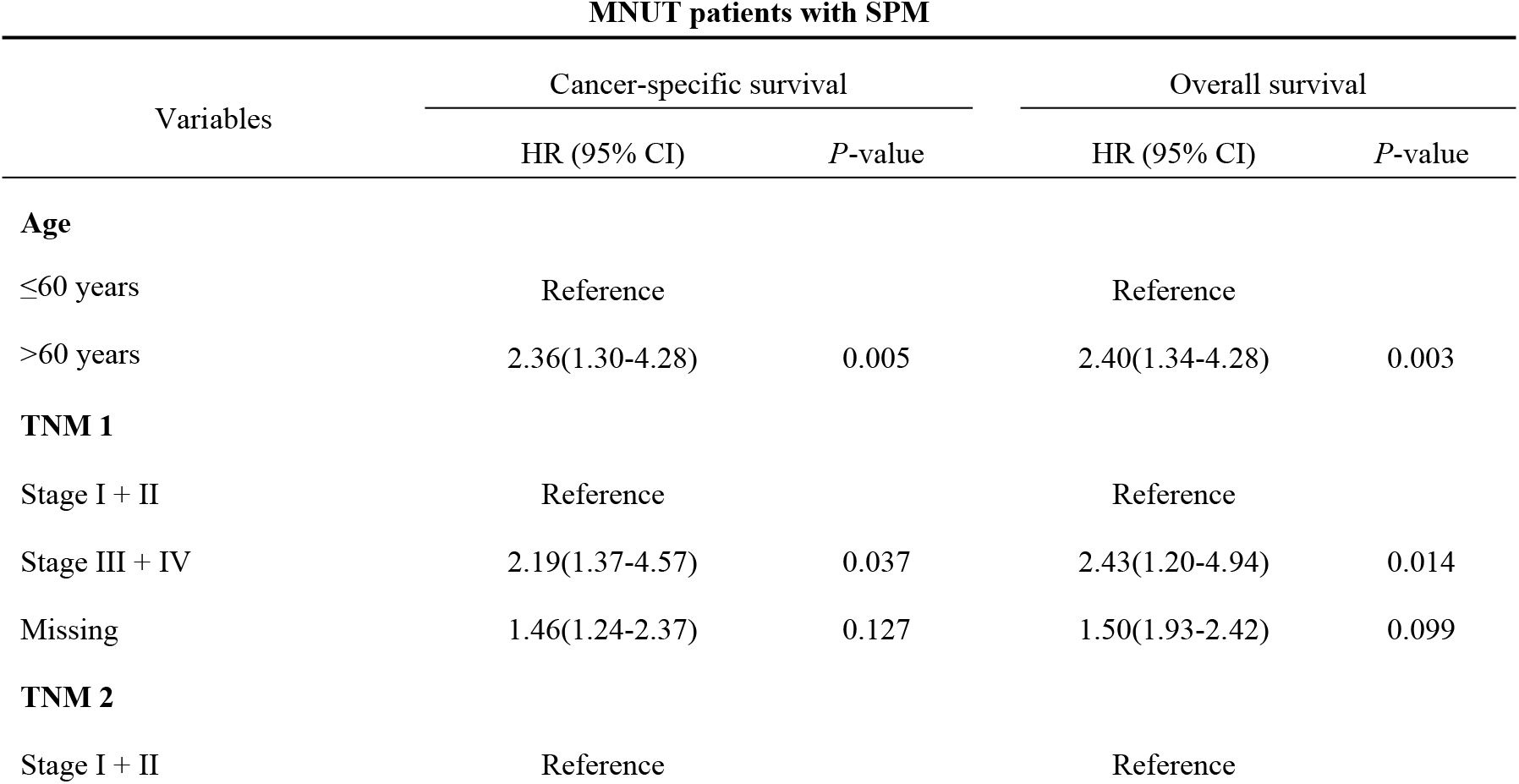
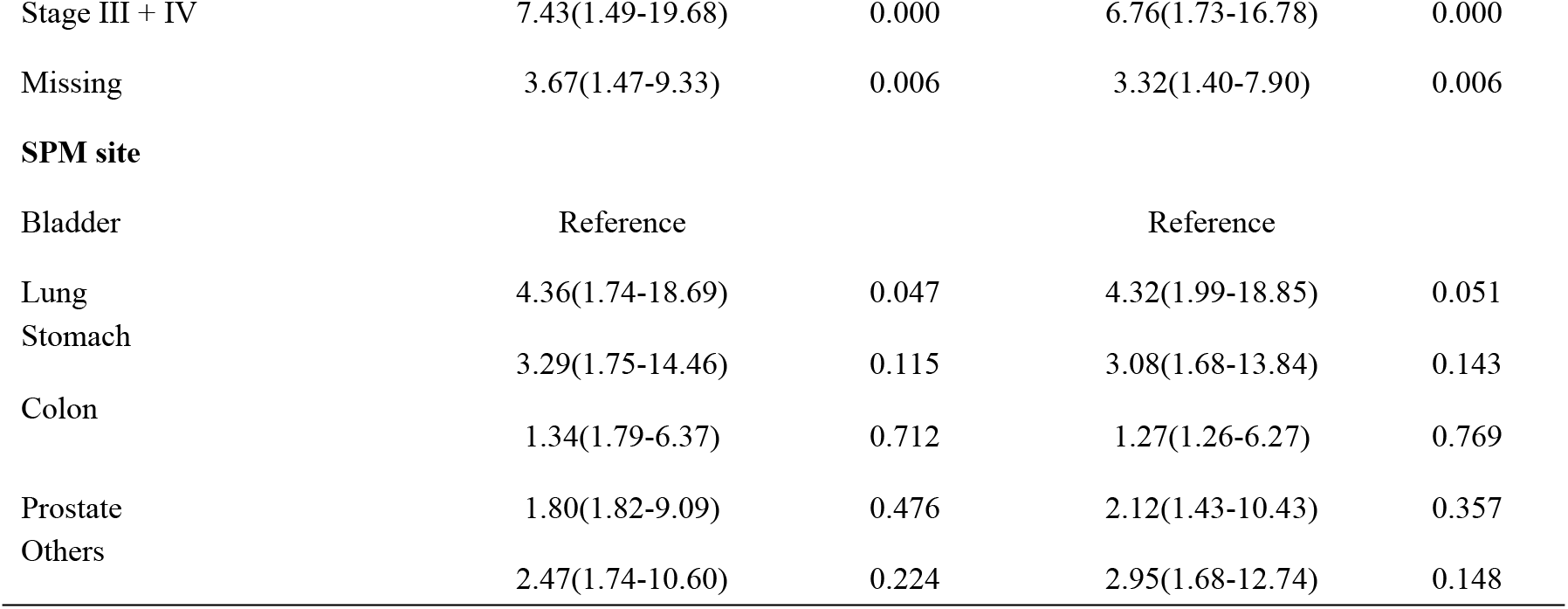
Multivariate Cox regression of independent risk factors for cancer-specific and overall survival in MNUT patients with SPM.

## Discussion

MNUTs are one of the most common types of malignant neoplasms. With the increase in the incidence of SPM, an increasing number of studies have begun to pay attention to the current status and risks of SPM(12). Although some studies have discussed the death risk of SPM in patients with specific urinary tract malignancies, such as kidney and prostate malignancies, due to a lack of data, the overall pattern of SPMs in MNUT and the risk factors related to survival still need to be further analysed and discussed.

In this study, we found that malignant neoplasms of the kidney and bladder were the most common MNUTs with SPM in Shanghai. This result was consistent with the research results of other scholars. For example, Jae Young Joung et al. found that the risk of SPM in patients with primary kidney cancer was higher than the cancer risk in the general population(13). In addition, this study found that the most common SPM site in patients with MNUT was the lung (35, 22.73%). This was consistent with the study findings of AMIT KHANAL(14). Because smoking is a well-known risk factor for malignant neoplasms of the lung, the main risk factors for malignant neoplasms of the bladder and other MNUTs include cigarette smoke, naphthylamine, azo dyes, and long-term use of cyclophosphamide or phenacetin(15, 16). Therefore, the increased risk of lung cancer in patients with a history of MNUT may be secondary to a common cause.

In addition, when analysing the combination of MNUTs, we observed a certain symbiosis of MNUTs, such as bladder+prostate and kidney+bladder. A similar phenomenon was found in previous studies. Following bladder cancer, the risk of renal pelvic, bladder, and ureter malignancies is significantly increased(14). This may be because the bladder, renal pelvis and ureter are lined by the same urothelium and are susceptible to the same risk factors for carcinogens in urine. Kotake T et al. found that the relative risk of prostate cancer was significantly increased after bladder cancer. The study analysed cases reported in the Japanese literature and found that the association with urinary system cancers may be because patients are highly sensitive to both cancers, or they may share similar carcinogenic processes, such as DNA repair and N-acetyltransferase polymorphism(17, 18).

According to Kaplan-Meier curve and multivariate Cox regression analyses, we found that patients in the advanced age group have a higher cancer death risk and all-cause death risk, and patients with stage III-IV diseases have a higher mortality rate when they are diagnosed with a first and second PM. This is mainly because patients diagnosed at a late stage cannot receive early treatment. Previous studies have suggested that the survival of patients with urinary tract malignancies with SPM depends to a large extent on the cancer stage at the time of diagnosis. For example, Jae Young Joung et al. found in a study that patients diagnosed with advanced renal cancer have poor survival outcomes. Early cancer is associated with a more favourable prognosis and a survival period of more than 10 years after complete resection(13). This means that if the public is encouraged to conduct early screening to identify urinary tract malignant tumours and their SPM earlier, they will be more likely to achieve good survival results.

Regarding the location of SPM, patients with urinary organ cancer with SPM in the lungs have a higher risk of death than patients with SPM in other locations. This is consistent with the results of previous studies. For example, Nicholas Donin MD and other scholars found in a study on the survival risk of cancer survivors with SPM in the United States that a large proportion of cancer survivors will develop and die from lung cancer(4). This result is also consistent with the known epidemiology of lung cancer. Lung cancer is still the leading cause of cancer-related deaths in many countries worldwide(1, 19-21).

In this study, factors such as sex and the time interval between the first and second PM did not have a significant effect on cancer mortality or all-cause mortality. These findings were different from those of previous studies, such as the Chinese Cancer Statistics Study, which showed that male cancer patients have a higher overall mortality rate(22). Of course, this conclusion needs to be confirmed by further studies with a larger sample size.

There are several limitations to consider. First, during the follow-up period, some patients lacked information on TNM staging, which led to a lack of information for these patients in the Cox regression analysis. Second, the sample size of this study was limited, and MNUT can only be used as a whole to analyse the impact of SPM on death. However, it is impossible to realize the analysis of risk factors for death of specific MNUTs combined with SPMs. In addition, due to the sample size, this study could not analyse 4 patients with 3 PMs in depth.

## Conclusions

The most common site of SPM is the lung. It is worth noting that patients with first and second PM in stages III-IV have a higher risk of cancer-specific death and all-cause death. Patients with MNUT whose SPM site is in the lung had a higher risk of specific and all-cause death than those with other sites of SPM. Future research is necessary to confirm our findings and translate them into SPM monitoring and treatment strategies to improve the prognosis of patients with MNUT after the development of SPM.

## Data Availability

The data are available on request from the corresponding author.

## Acknowledgments

Conceptualization, Xiang Gao and Jianwei Shi; methodology, Zhaoxin Wang and Hua Jin; software, Hui Cao and Liang Zhou; validation, Dehua Yu, and Jinsong Geng; formal analysis, Yongbin Zou, Yipeng Lv, Wenya Yu and Yichen Chen; resources, Xiaopan Li and Hua Jin; data curation, Xiang Gao and Haiyi Jia; writing—original draft preparation, Xiang Gao and Haiyi Jia; writing—review and editing, Xiang Gao and Jianwei Shi; project administration, Xiang Gao and Jianwei Shi. All authors have read and agreed to the published version of the manuscript.

## Disclosure

### Ethics statement

Ethical approval for the research protocol was provided by the Medical Research Ethics Committee of the School of Public Health, Fudan University (IRB#2016-04-0586). Information was de-identified prior to analysis. The guidelines outlined in the Declaration of Helsinki were followed.

### Registry and the Registration No. of the study/trial

N/A.

### Informed Consent

N/A.

### Animal Studies

N/A.

## References

1. Sung H, Ferlay J, Siegel RL, Laversanne M, Soerjomataram I, Jemal A, et al. Global cancer statistics 2020: GLOBOCAN estimates of incidence and mortality worldwide for 36 cancers in 185 countries. Ca-a Cancer Journal for Clinicians. 2021;71(3):209–49.

2. Wang Z, Yin Y, Wang J, Zhu Y, Li X, Zeng X. Standardized Incidence Rate, Risk and Survival Outcomes of Second Primary Malignancy Among Renal Cell Carcinoma Survivors: A Nested Case-Control Study. Frontiers in Oncology. 2021;1.

3. Zheng G, Sundquist K, Sundquist J, Forsti A, Hemminki O, Hemminki K. Bladder and upper urinary tract cancers as first and second primary cancers. Cancer Reports. 2021.

4. Donin N, Filson C, Drakaki A, Tan H-J, Castillo A, Kwan L, et al. Risk of Second Primary Malignancies Among Cancer Survivors in the United States, 1992 Through 2008. Cancer. 2016;122(19):3075–86.

5. Beisland C, Talleraas O, Bakke A, Norstein J. Multiple primary malignancies in patients with renal cell carcinoma: a national population-based cohort study. Bju International. 2006;97(4):698–702.

6. Liu Y, Zhang P, Zhang Y, Zheng L, Xu W, Hou D, et al. Clinical characteristics and overall survival nomogram of second primary malignancies after prostate cancer, a SEER population-based study. Scientific Reports. 2021;11(1).

7. Koo KC, Yoo H, Kim KH, Park SU, Han KS, Rha KH, et al. Prognostic Impact of Synchronous Second Primary Malignancies on the Overall Survival of Patients with Metastatic Prostate Cancer. Journal of Urology. 2015;193(4):1239–44.

8. Deng L, Hardardottir H, Song H, Xiao Z, Jiang C, Wang Q, et al. Mortality of lung cancer as a second primary malignancy: A population-based cohort study. Cancer Medicine. 2019;8(6):3269–77.

9. Khanal A, Lashari BH, Kruthiventi S, Arjyal L, Bista A, Rimal P, et al. The risk of second primary malignancy in patients with stage Ia non-small cell lung cancer: a US population-based study. Acta Oncologica. 2018;57(2):239–43.

10. Edge SB, Compton CC. The American Joint Committee on Cancer: the 7th Edition of the AJCC Cancer Staging Manual and the Future of TNM. Annals of Surgical Oncology. 2010;17(6):1471–4.

11. Goldfarb M, Rosenberg AS, Li Q, Keegan THM. Impact of Latency Time on Survival for Adolescents and Young Adults With a Second Primary Malignancy. Cancer. 2018;124(6):1260–8.

12. Rashed WM, Saad A, Al-Husseini M, Galal AM, Ismael AM, Al-Tayep AM, et al. Incidence of adrenal gland tumor as a second primary malignancy: SEER-based study. Endocrine Connections. 2018;7(10):1040–8.

13. Joung JY, Kwon W-A, Lim J, Oh C-M, Jung K-W, Kim SH, et al. Second Primary Cancer Risk among Kidney Cancer Patients in Korea: A Population-Based Cohort Study. Cancer Research and Treatment. 2018;50(1):293–301.

14. Khanal A, Budhathoki N, Singh VP, Shah BK. Second Primary Malignancy in Bladder Carcinoma-A Population-based Study. Anticancer Research. 2017;37(4):2033–6.

15. Dawson C, Whitfield H. ABC of urology - Urological malignancy .1. Prostate cancer. British Medical Journal. 1996;312(7037):1032–4.

16. Salminen E, Pukkala E, Teppo L. BLADDER-CANCER AND THE RISK OF SMOKING-RELATED CANCERS DURING FOLLOW-UP. Journal of Urology. 1994;152(5):1420–3.

17. Kotake T, Kiyohara H. MULTIPLE PRIMARY CANCERS (MPC) ASSOCIATED WITH BLADDER-CANCER - AN ANALYSIS OF THE CLINICAL AND AUTOPSY CASES IN JAPAN. Japanese Journal of Clinical Oncology. 1985;15:201–10.

18. Kinoshita Y, Singh A, Rovito PM, Jr., Wang CY, Haas GP. Double primary cancers of the prostate and bladder: a literature review. Clinical prostate cancer. 2004;3(2):83–6.

19. Cao W, Chen H-D, Yu Y-W, Li N, Chen W-Q. Changing profiles of cancer burden worldwide and in China: a secondary analysis of the global cancer statistics 2020. Chinese Medical Journal. 2021;134(7):783–91.

20. Ferlay J, Colombet M, Soerjomataram I, Parkin DM, Pineros M, Znaor A, et al. Cancer statistics for the year 2020: An overview. International Journal of Cancer. 2021;149(4):778–89.

21. Canadian Cancer Statistics: A 2020 special report on lung cancer. Health Promotion and Chronic Disease Prevention in Canada-Research Policy and Practice. 2020;40(10):325-.

22. Feng R-M, Zong Y-N, Cao S-M, Xu R-H. Current cancer situation in China: good or bad news from the 2018 Global Cancer Statistics? Cancer Communications. 2019;39.

